# TreatmentPatterns: An R package to analyze treatment patterns of a study population of interest

**DOI:** 10.1101/2022.01.24.22269588

**Authors:** Aniek F. Markus, Katia M.C. Verhamme, Jan A. Kors, Peter R. Rijnbeek

## Abstract

**Background and objectives:** There is an increasing interest to use real-world data to illustrate how patients with specific medical conditions are treated in real life. Insight in the current treatment practices helps to improve and tailor patient care. We aim to provide an easy tool to support the development and analysis of treatment pathways for a wide variety of medical conditions.

**Methods:** We formally defined the process of constructing treatment pathways and developed an open-source R package TreatmentPatterns (https://github.com/mi-erasmusmc/TreatmentPatterns) to enable a reproducible and timely analysis of treatment patterns.

**Results:** The developed package supports the analysis of treatment patterns of specific populations of interest. We demonstrate the functionality of the package by analyzing the treatment patterns of three common chronic diseases (type II diabetes mellitus, hypertension, and depression) in the Dutch Integrated Primary Care Information (IPCI) database.

**Conclusion:** TreatmentPatterns is a tool to make the analysis of treatment patterns more accessible, more standardized, and more interpretation friendly. This tool can facilitate and contribute to the accumulation of knowledge on real-world treatment patterns across disease domains. We encourage researchers to further adjust and add custom analysis to the package based on their research needs.

## BACKGROUND

There is an increasing interest to use real-world data to illustrate how patients with specific medical conditions are treated in real life. Real-world data is already used for post-market safety surveillance of medicines, to monitor drug consumption and related costs, to detect inappropriate prescribing and to assess compliance with treatment guidelines [1]. This research focusing on treatment patterns allows to analyze drug utilization beyond the volume of drug uptake (i.e. incidence and prevalence numbers on prescribing/dispensing), as it allows to investigate treatment pathways (i.e. first-line and subsequent treatments over time). The latter is crucial to investigate adherence to treatment guidelines in their entirety. Clinical guidelines are systematically developed recommendations informed by a review of scientific evidence and an assessment of the benefits and harms of alternative treatments for a specific medical condition [2]. These guidelines therefore support evidence-based medicine by serving as a ‘best practice’. Unfortunately, clinical guidelines are often not well adhered to in practice for a variety of reasons including lack of applicability, organizational constraints, or lack of awareness [3].

Knowledge of the current treatment practices is an important starting point to improve clinical practice and from a community perspective to facilitate the rational use of drugs. Evidence from real-world data has driven changes in clinical guidelines, but it is recognized that this data is not yet used to its full potential [1]. An increasing amount of routine health care data is available in the form of electronical health records, claims data and registries, which allows the analysis of treatment pathways in real life [4-7]. Currently, there is no easy-to-use tool available that is still flexible enough to accommodate specific research needs. Barriers to analyze treatment practices might be a lack of data interoperability and a high-level of required resources (i.e. amount of time and level of expertise).

In this paper we present TreatmentPatterns, an open-source R package to enable the analysis of treatment patterns that overcomes these barriers. The Observational Medical Outcomes Partnership (OMOP) common data model (CDM) maps data in different structures, formats and terminologies to one common standard to promote interoperability of health care data [8]. This allows to perform multi-database studies without the need to share patient-level data. Moreover, the OMOP CDM allows for the development of standardized tools, which is important to produce reproducible and timely real-world evidence. A wide range of tools is already available to support research on the OMOP CDM; Health Analytics Data-to-Evidence Suite (HADES)^1^ is a set of open source R packages for large-scale analytics, including population characterization, population-level causal effect estimation, and patient-level prediction. It also provides supporting packages for new analytical pipelines. These building blocks are used by TreatmentPatterns to analyze treatment patterns of a study population of interest. The package can be executed against the OMOP CDM, but the main parts of the package are also usable with other data formats.

## OBJECTIVES

We aimed to provide a user-friendly tool to support the development and analysis of treatment pathways for a wide variety of medical conditions. By making the analysis of treatment patterns more accessible, more standardized, and more interpretation friendly, we aim to contribute to the accumulation of knowledge on real-world treatment patterns across disease domains. We demonstrate the package by analyzing the treatment patterns of three common chronic diseases (type II diabetes mellitus, hypertension, and depression) using data from the Dutch Integrated Primary Care Information (IPCI) database^2^.

## METHODS

In this section, we first formally define the problem of constructing treatment pathways following earlier work [4]. Based on the problem formalization, we then describe the implementation of the open-source R package TreatmentPatterns, which enables a reproducible and timely analysis of treatment patterns.

### Problem formalization

We defined the problem as follows: for a specified study population (i.e. target cohort), the goal is to find the treatment pathway consisting of selected “treatments of interest” (i.e. event cohorts). Treatments of interest (i.e. events) can be drug prescribing and/or dispensing, but could as well be other therapies, procedures, or measurements. The treatment pathway is defined as the sequence of treatments of interest over time. The target cohort is the set of patients for whom we want to study treatment pathways and should be defined by a clear set of inclusion/exclusion criteria. Similarly, event cohorts specify which patients have received a given treatment of interest. We use the term event era to refer to the span of time a patient is exposed to a specific treatment of interest. How to define target and event cohorts is elaborated on in other sources (e.g. Chapter 10 ‘Defining cohorts’ in the Book Of OHDSI [9]). The challenge addressed in this paper is how the treatments of interest should be processed to construct treatment pathways.

For each individual in a target cohort, we extract the treatments from his or her medical file after the index date. Note that individuals can have many different, sometimes simultaneously occurring treatments in their medical file. The index date is the moment in time the individual enters the target cohort. Figure 1 shows an example medical file for an individual receiving treatments A, B and C. The figure visualizes the decisions that need to be made to construct the treatment pathway for this individual.

**Figure 1:**
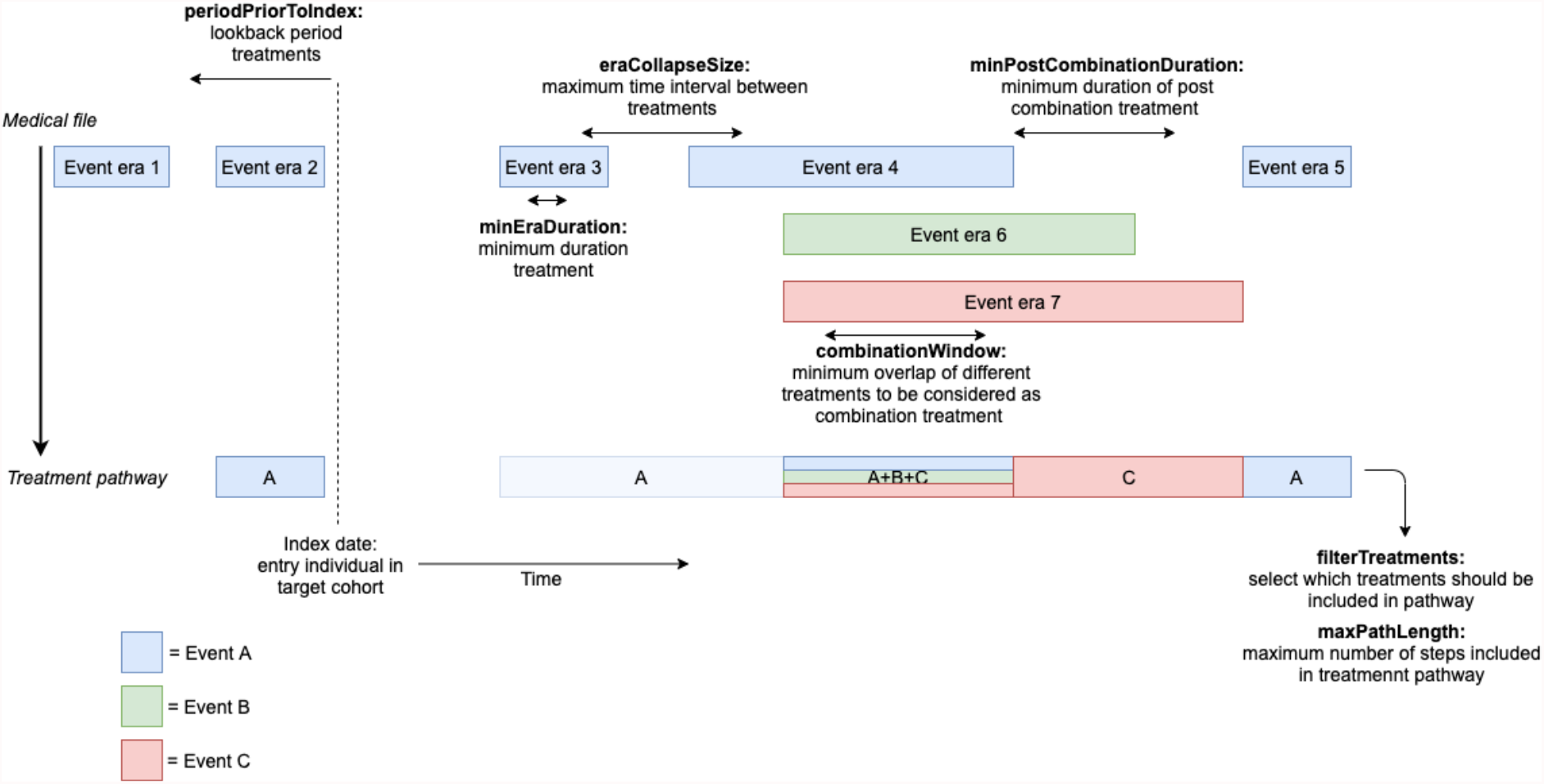
Summary of decisions to construct individual treatment pathways. The events A, B, and C refer to the treatments of interest. For full detailed steps from medical file to treatment pathway see Appendix A.

To create treatment pathways, the following decisions need to be made:

#### 1. Lookback period treatments (periodPriorToIndex)

The first decision that needs to be made is the moment in time from which selected treatments of interest should be included in the treatment pathway. The default is all treatments starting after the index date of the target cohort. For example, for a target cohort consisting of newly diagnosed patients, treatments after the moment of first diagnosis are included. However, it might also be desirable to include (some) treatments prior to the index date, for instance in case a specific disease diagnosis is only confirmed after initiating treatment. Therefore, *periodPriorToIndex* specifies the period (i.e. number of days) prior to the index date from which treatments should be included.

#### 2. Minimum duration treatment (minEraDuration)

The duration of extracted event eras may vary a lot and it can be preferable to limit to only treatments exceeding a minimum duration. Hence, *minEraDuration* specifies the minimum time an event era should last to be included in the analysis.

#### 3. Maximum time interval between treatments (eraCollapseSize)

If an individual receives the same treatment for a longer period of time (e.g. need of chronic treatment), one is likely to need refills. As patients are not 100% adherent, there might be a gap between two subsequent event eras. Usually, these eras are still considered as one treatment episode and the *eraCollapseSize* defines the maximum gap within which two eras of the same event cohort would be collapsed into one era (i.e. seen as continuous treatment instead of a stop and re-initiation of the same treatment).

#### 4. Minimum overlap of different treatments to be considered as combination treatment (combinationWindow)

Patients often receive different treatments at the same time, which may occur with full overlap or partial overlap. If the period of overlap is short, this is defined as a *switch* in treatment, which means that the earlier treatment is switched to a new treatment. If the overlap is longer, we assume that both treatments are received at the same time. To differentiate between the two options, *combinationWindow* specifies the time that two event eras need to overlap to be considered a combination treatment. If there are more than two overlapping event eras, we sequentially combine treatments, starting from the first two overlapping event eras.

#### 5. Minimum duration of post combination treatment (minPostCombinationDuration)

As a result of creating combination treatments, there might be short remaining periods of a single treatment before or after the combination treatments. For similar reasons as described above, we might only want to consider treatments if they last for a sufficient period of time. Therefore, *minPostCombinationDuration* defines the minimum time that an event era before or after a generated combination treatment should last to be included in the pathway as a separate treatment.

#### 6. Select which treatments should be included in pathway (filterTreatments)

Depending on the research question, we might be interested to include repeated treatments in the pathway or not. The *filterTreatments* parameter allows to specify whether to include all treatments (‘All’; A – A – A+B+C – C – A), to remove sequential repeated treatments (‘Changes’; A – A+B+C – C – A), or to only include first time occurrences of treatments (‘First’; A – A+B+C – C).

#### 7. Maximum number of treatments included in pathway (maxPathLength)

Finally, *maxPathLength* specifies the maximum number of treatments included in the treatment pathway (i.e. the number of layers in a sunburst plot).

### Implementation

The R package TreatmentPatterns implements the problem formalization described above and is structured as shown in Figure 2. The package allows to run the full analysis at once with the *executeTreatmentPatterns* function or by sequentially running the individual subfunctions (i.e. *createCohorts, cohortCharacterization* (optional for databases mapped to the OMOP CDM), *constructPathways, generateOutput*, and *launchResultsExplorer*).

**Figure 2:**
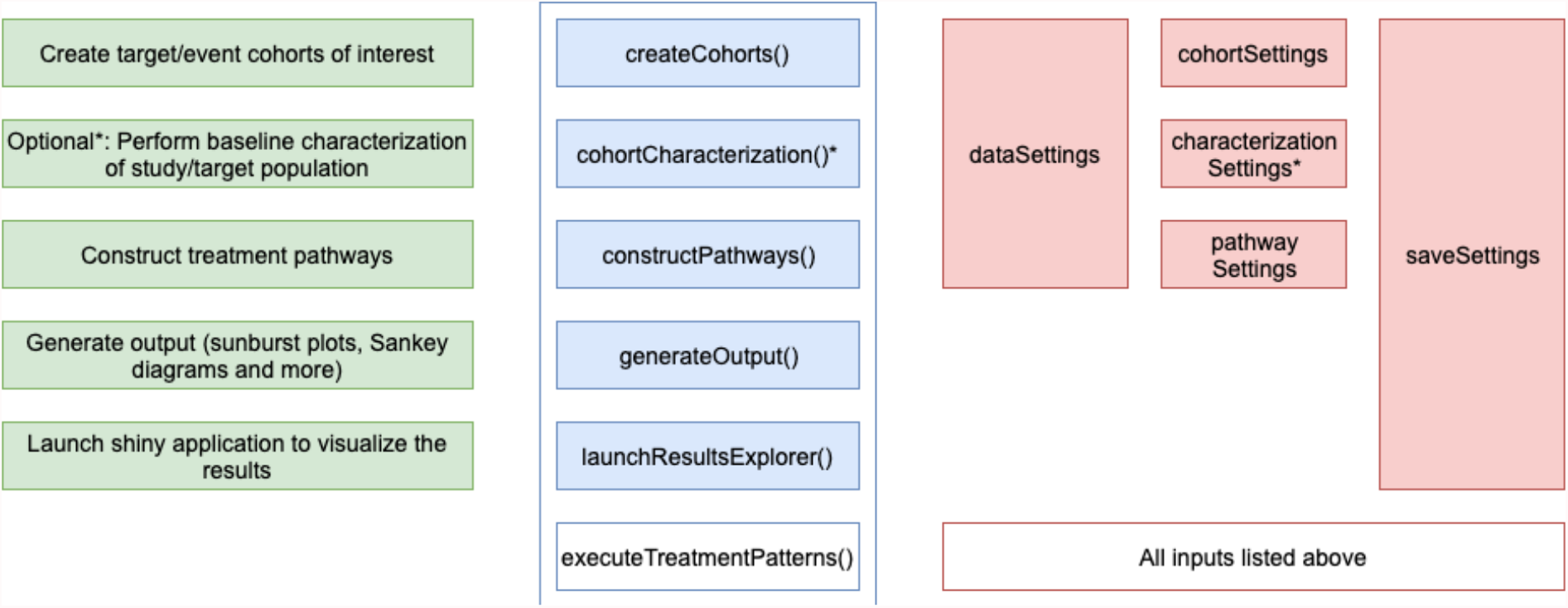
Structure of R package TreatmentPatterns. * Only for data in OMOP CDM format.

The main inputs of the package are the target and event cohorts of interest, which can be cohort definitions to allow automatic cohort extraction from databases mapped to the OMOP CDM or a csv file to directly import existing target and event cohorts (to be specified in *cohortSettings*). The package also allows to investigate baseline characterization of the study population including a wide range of pre-specified covariates (e.g. age/gender, Charlson comorbidity index score, comorbid conditions with different look-back periods), which can be specified in *characterizationSettings*. Moreover, it is possible to add custom covariates using SQL code. Furthermore, the *pathwaySettings* for the construction of treatment pathways need to be specified. In addition to the settings related to the creation of the individual treatment pathways above, these include output settings to aggregate the treatment pathways (e.g. *minCellCount, groupCombinations*). For a list of pathway settings with description and expected input we refer to Appendix B. Other necessary inputs are *dataSettings* and *saveSettings*, which indicate the location of the input data and output respectively.

After cohort extraction (and optional baseline characterization of the study population), the individual treatment pathway for each patient is constructed as follows:

1. Pre-process the medical file according to, amongst others, *periodPriorToIndex, minEraDuration*, and *eraCollapseSize*.
2. Identify which event eras have overlap for each person and select the first two overlapping event eras of different events per person.
3. We now distinguish three cases of overlap (see Figure 3):
  a. If the overlap of the two event eras is smaller than *combinationWindow* AND the overlap is not equal to the total length of one of the event eras, then this is considered a *Switch*. The end date of the earlier event will be adapted to the start of the later event. If the overlap of the two event eras is at least *combinationWindow* OR the overlap is equal to the total length of one of the events, this is considered:
  b. *FRFS* (‘First Received, First Stopped’) if the end date of the earlier starting event era is before or at the end date of the later starting event era.
  c. *LRFS* (‘Last Received, First Stopped’) if the end date of the earlier starting event era is after the end date of the later starting event era.
4. Filter the event eras before or after a generated combination treatment based on *minPostCombinationDuration* and go back to Step 2. Stop if there are no more event eras with overlap.

**Figure 3:**
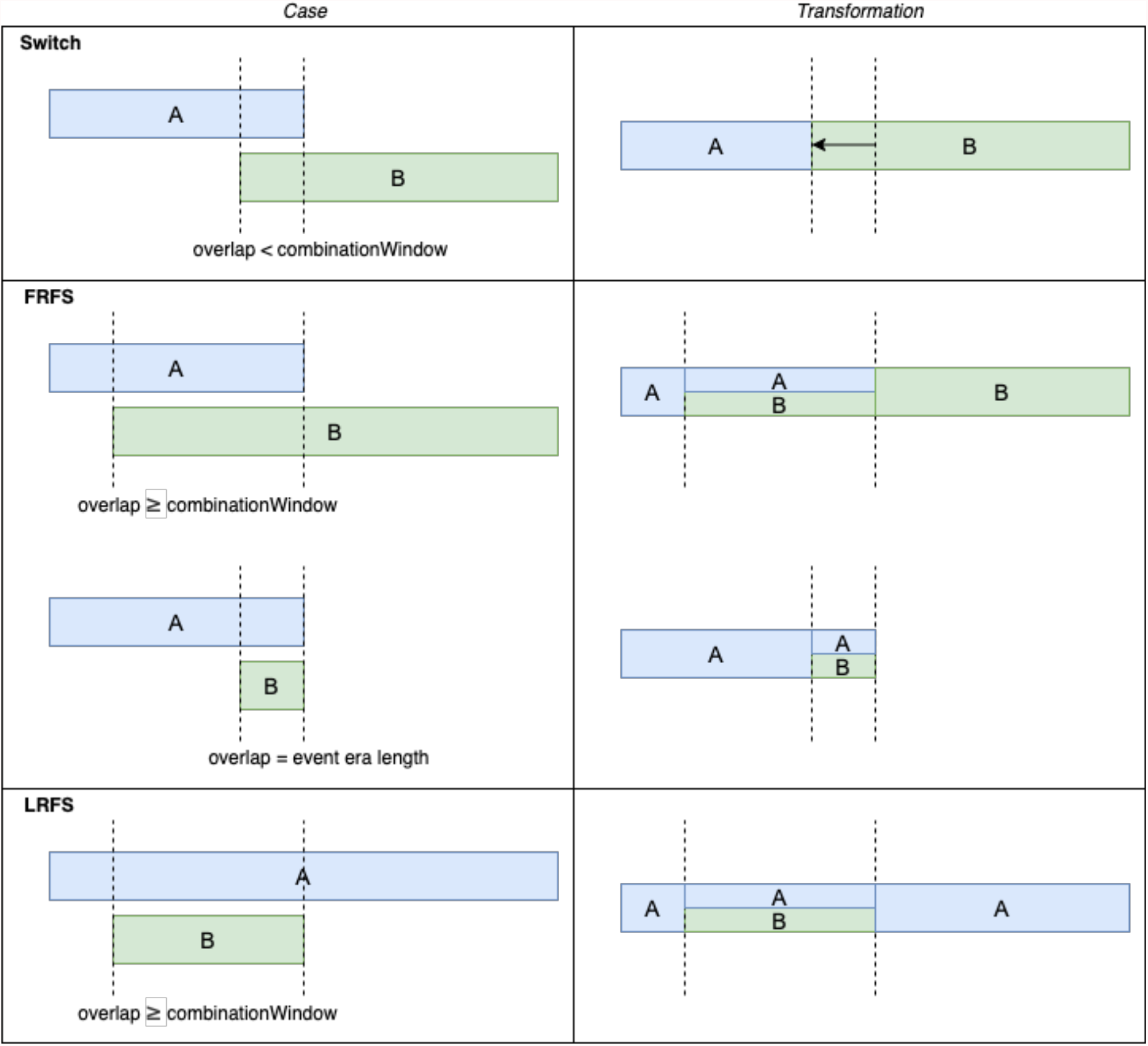
Three cases of overlap can occur: a) *Switch*, b) *FRFS* (‘First Received, First Stopped’), and c) *LRFS* (‘Last Received, First Stopped’).

After constructing the individual treatment pathways, the results are aggregated for each target cohort. The package provides several functions for displaying the results of the treatment patterns analysis in both graphical and tabular formats. The package creates sunburst plots, Sankey diagrams, and various other outputs (e.g. percentage of people treated, treatment changes over time, average duration of event eras) to give insight in first-line, second-line, and higher-line treatments. *Sunburst plots* of treatment patterns show the first treatment in the center and subsequent treatments in the surrounding outer layers. Each color represents a treatment (as defined by the event cohorts) and a layer with multiple colors indicates a combination therapy. This shows the full treatment pathway of patients over time. *Sankey diagrams* also visualize treatment patterns, but show follow-up treatment conditional on the previously received treatment by directed arrows from left to right. The size of the arrows is proportional to the number of patients. All results can be explored in an interactive Shiny application.

For full details and instructions to use the package (including a vignette and package manual) we refer to GitHub: https://github.com/mi-erasmusmc/TreatmentPatterns.

## RESULTS

To demonstrate the functionality of the package, we present an example study analyzing treatment patterns of three common chronic diseases using data from the Dutch IPCI database. The IPCI database contains records of more than two million patients enrolled at selected general practitioners (GPs) throughout the Netherlands [10]. We aimed to study treatment patterns of patients with type II diabetes mellitus, hypertension, and depression.

### Step 1: define target/event cohorts

We created three target cohorts of patients with index date at first diagnosis of the respective disease. Patients needed to have at least 365 days prior observation database time and 1,095 days follow-up time after index date (to allow sufficient time to investigate treatment patterns). We only included adult patients with a first diagnosis since 2010. The event cohorts include frequently prescribed drugs for each of the chronic diseases. An overview of the study is provided in Table 1, for more details and code lists we refer to Appendix C.

**Table 1:**
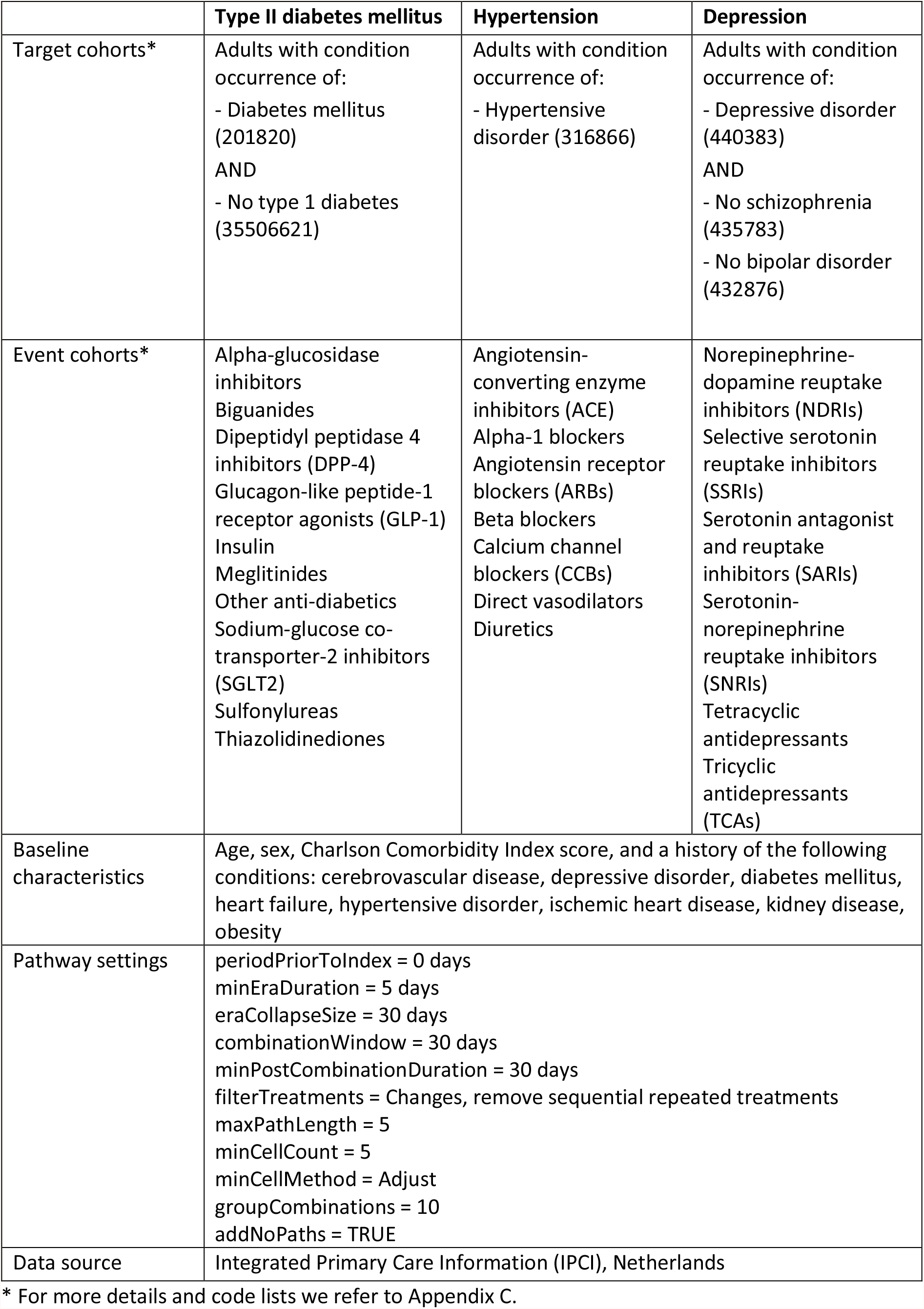
Overview of example study on type II diabetes mellitus, hypertension, and depression.

### Step 2: specify baseline characteristics of interest

We chose to include the following baseline characteristics of interest: age, sex, Charlson Comorbidity Index score, and a history of the following conditions: cerebrovascular disease, depressive disorder, diabetes mellitus, heart failure, hypertensive disorder, ischemic heart disease, kidney disease, obesity. For each target cohort we excluded the history of the corresponding disease from the list of baseline characteristics.

### Step 3: specify settings to construct treatment pathways

We constructed treatment pathways from the moment of first diagnosis, including the treatments of interest as specified for the different study populations in Table 1. We restricted the analysis to event eras that last at least 5 days (*minEraDuration*) and considered a treatment to be a combination if the overlap is 30 days (combinationWindow). We included all changes between treatments (*filterTreatments*), thereby removing sequential repeated treatments but keeping recurring treatments in the pathways (i.e. A – A+B+C – C – A). For the full list of pathway settings see Table 1.

### Step 4: execute study

The complete treatment patterns study was executed by running the *executeTreatmentPatterns* function after defining the setting objects (see Appendix D for a code example). This set up has the advantage that limited programming expertise is required as the package will automatically execute all steps and produce an interactive Shiny application, the results of which are subsequently discussed.

### Step 5: check out results

The results of the example study are available at: https://aniekmarkus.shinyapps.io/TreatmentPatterns/. We found 50,285 type II diabetes mellitus patients, 120,675 hypertension patients, and 32,567 patients with depression. Baseline characteristics showed that the proportion of males is slightly lower than that of females in patients newly diagnosed with hypertension (44.2%) and depression (35.8%) whereas for type II diabetes mellitus no female predominance was observed. The interface allows to easily compare baseline characteristics across databases or study populations (as shown in Figure 4).

**Figure 4:**
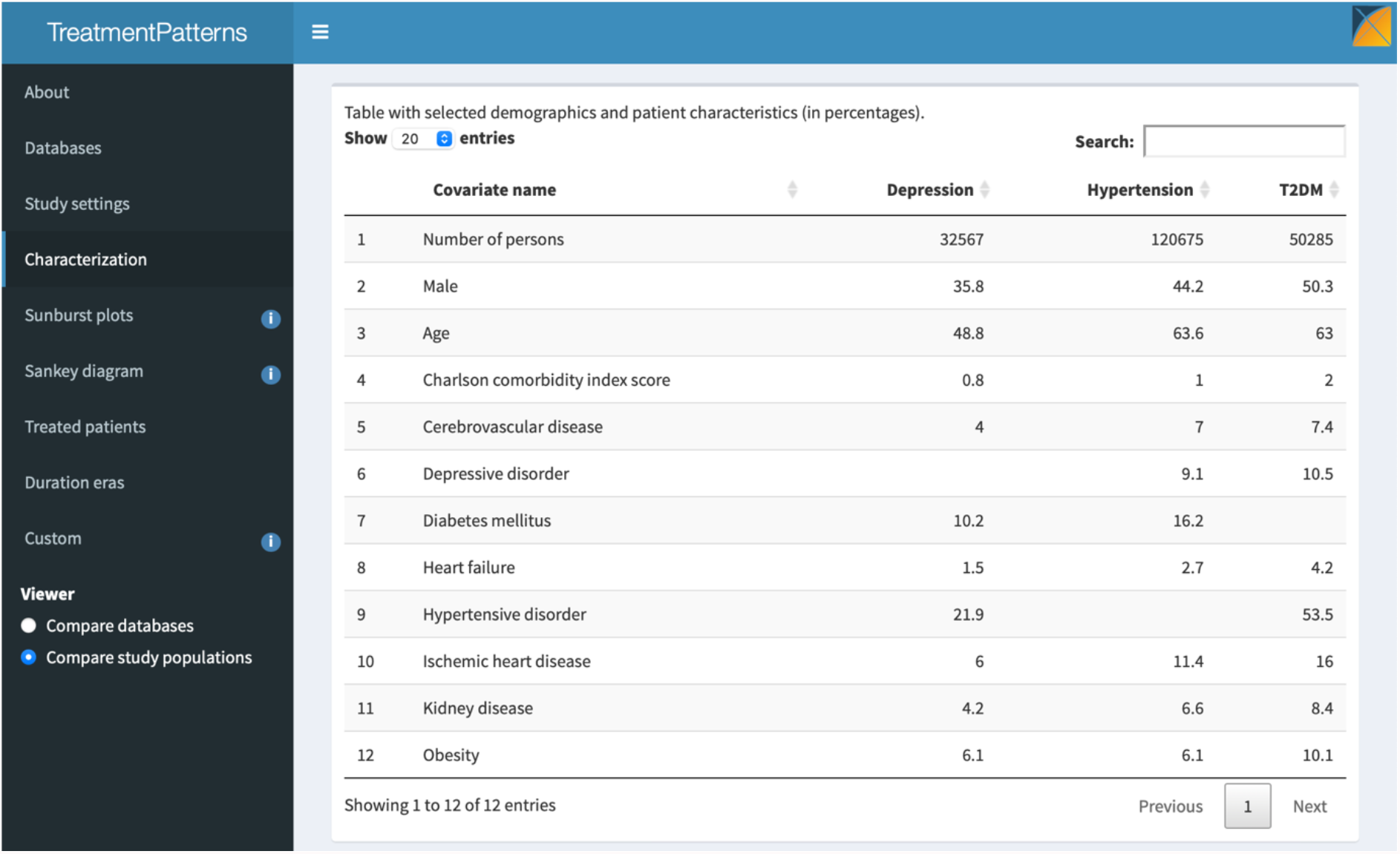
Baseline characterization of study populations in IPCI.

Treatment pathways were visualized in the form of various plots and an example of a sunburst plot is shown in Figure 5 (for remaining plots see Appendix E). For type II diabetes mellitus, biguanides were the most frequently prescribed first-line treatments (62.4% of treated patients) followed by insulin (9.9% of treated patients). GPs thus seems to align with regard to the type of first-line treatment for type II diabetes mellitus. The proportion of switching was low as 35.2% of treated patients remained on biguanides (i.e. did not receive another type of treatment afterwards). More variability in the type of second-line treatments was observed, and this also included more combination treatments (62.3% as opposed to 11.1%). For hypertension, ACE inhibitors (26.7% of treated patients), beta blockers (19.1% of treated patients), CCBs (17.1% of treated patients) and ARBs (16.5% of treated patients) were all frequently prescribed as first treatment (see Figure 1 in Appendix E). For depression, SSRIs were the most frequently prescribed first-line treatment (61.2% of treated patients), followed by three types of medication that were each received by at least 10% of the treated patients: SNRIs, TCAs, and tetracyclic antidepressants (see Figure 2 Appendix E). Interestingly, patients treated for depression often did not proceed to a second-line treatment (only 24.8%). Use of combination treatments as first-line treatment was mainly observed in patients with hypertension (15.0%) and type II diabetes mellitus (11.1%), but was rare in patients with depression (1.9%).

**Figure 5:**
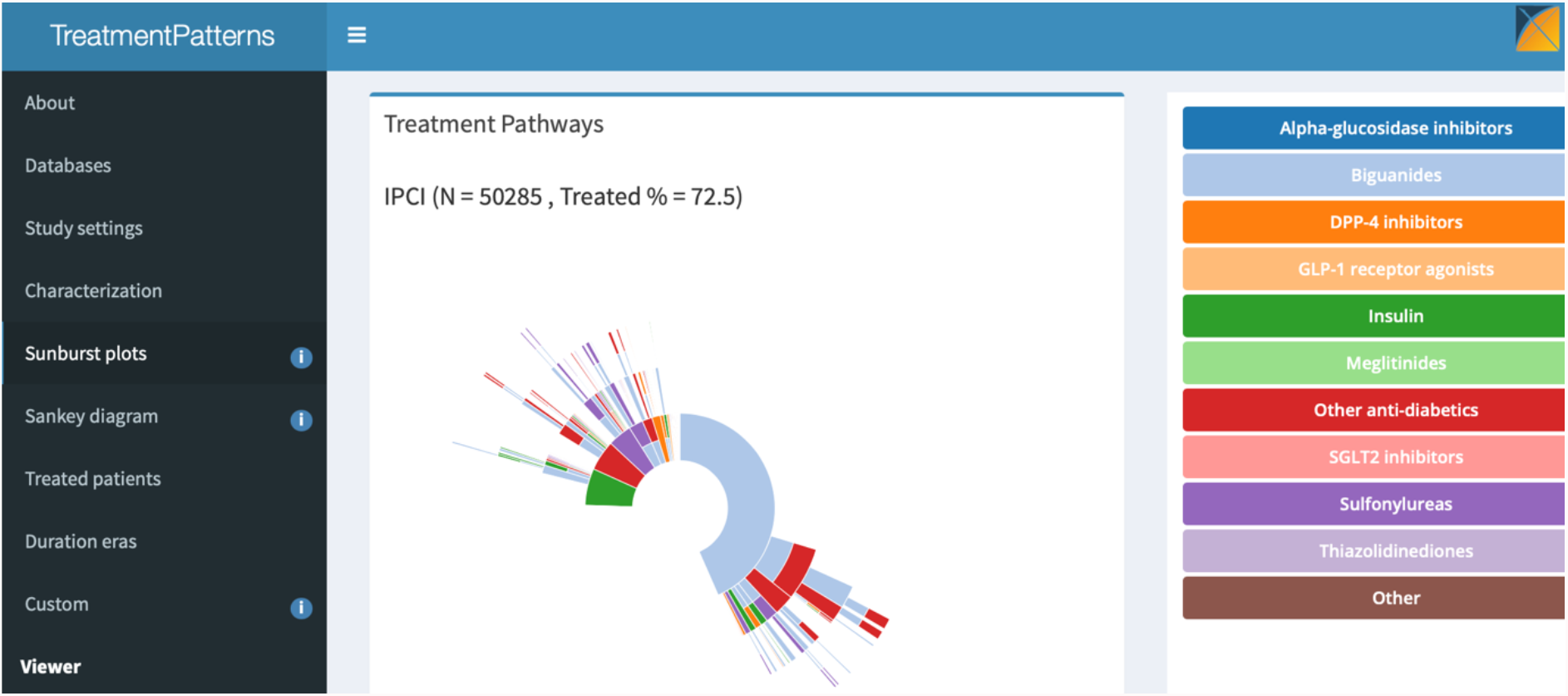
Sunburst plot visualizing the treatment pathways of patients with type II diabetes mellitus in IPCI.

Finally, the heatmaps indicating the duration of event eras (see Figures 6-8 in Appendix E) showed that the mean duration of first-line treatment for type II diabetes mellitus was 661 days, for hypertension 674 days, and for depression 658 days.

## DISCUSSION

In this paper, we formalize the construction of treatment pathways and present an R package implementing the presented analysis. This allows researchers to give insight in treatment patterns of a specified study population (target cohort) and selected treatments of interest (event cohorts) in a reproducible and timely manner. TreatmentPatterns facilitates the execution of these studies at a large-scale, i.e. across data sources, countries, and disease domains, while allowing to easily adjust settings and perform sensitivity analysis to investigate the robustness of the results (both within and between data sources).

Insight in the current treatment practices helps to improve clinical practice. It gives insight in the choice of treatment and preferences of doctors taking into account patient’s phenotype characteristics. In addition, it provides indirect information on pharmaceutical expenditure and potential health care budget constraints. Furthermore, this provides knowledge on the extent to which clinical guidelines are adhered to, which is an important step in further implementing and/or revisiting current guidelines.

Previous work analyzing treatment pathways is often addressing specific use cases and includes for example the assessment of current treatment practices of patients with type II diabetes mellitus [4 7], hypertension [4], depression [4 5], and epilepsy [6]. Some methodological work on treatment patterns exists, but focuses for example on extracting treatment patterns using a multi-view similarity fusion method [10] or alternative visualization techniques [11]. To our knowledge, no earlier work formally defined the process of constructing treatment pathways from medical files. The already existing ‘Cohort Pathways’ tool^3^, which was used for some of the before-mentioned studies, has the disadvantage that it has a very limited number of settings and is not customizable to address specific research needs. The presented R package is thus an addition to the existing base of methods available to generate evidence from routinely collected data.

TreatmentPatterns is a user-friendly tool to support the development and analysis of treatment pathways without requiring much programming. Therefore, the target audience is large and includes - but is not limited to - researchers, clinicians, the pharmaceutical industry, regulatory bodies, and health technology assessment agencies. The quality of the analysis is dependent on the target and event cohort definitions (i.e. phenotypes), which need to be created and validated by the user (e.g. using the OHDSI CohortDiagnostics tool). However, this is a limitation related to the quality of the data being used that holds for all types of observational data studies and is not specific for research investigating treatment patterns. Finally, it should be noted that the analysis of treatment patterns in observational data is limited to prescribing and/or dispensing data. Hence, it is not possible to infer actual treatment intake. This should be taken into account when interpreting the results.

In conclusion, this tool is intended to make the analysis of treatment patterns more accessible, more standardized, and more interpretation friendly. We hope it thereby contributes to the accumulation of knowledge on real-world treatment patterns across disease domains. Note that there might be disease-specific features of interest, that are not included in the current package. We thus encourage researchers to further adjust and add custom analysis to the package based on their research needs.

## Data Availability

All data produced in the present study are available upon reasonable request to the authors

## ACKNOWLEDGEMENTS

The authors would like to thank the Observational Health Data Sciences and Informatics (OHDSI) community for building the foundation necessary to perform large-scale, international, multi-database studies through the development of the OMOP CDM and open-source tools. In particular, we like to thank the authors of the earlier paper ‘Characterizing treatment pathways at scale using the OHDSI network’ [4], which has provided the initial foundation and rationale to develop the TreatmentPatterns package.

## SUPPLEMENTARY MATERIALS

### Appendix A: Example to construct individual treatment pathway

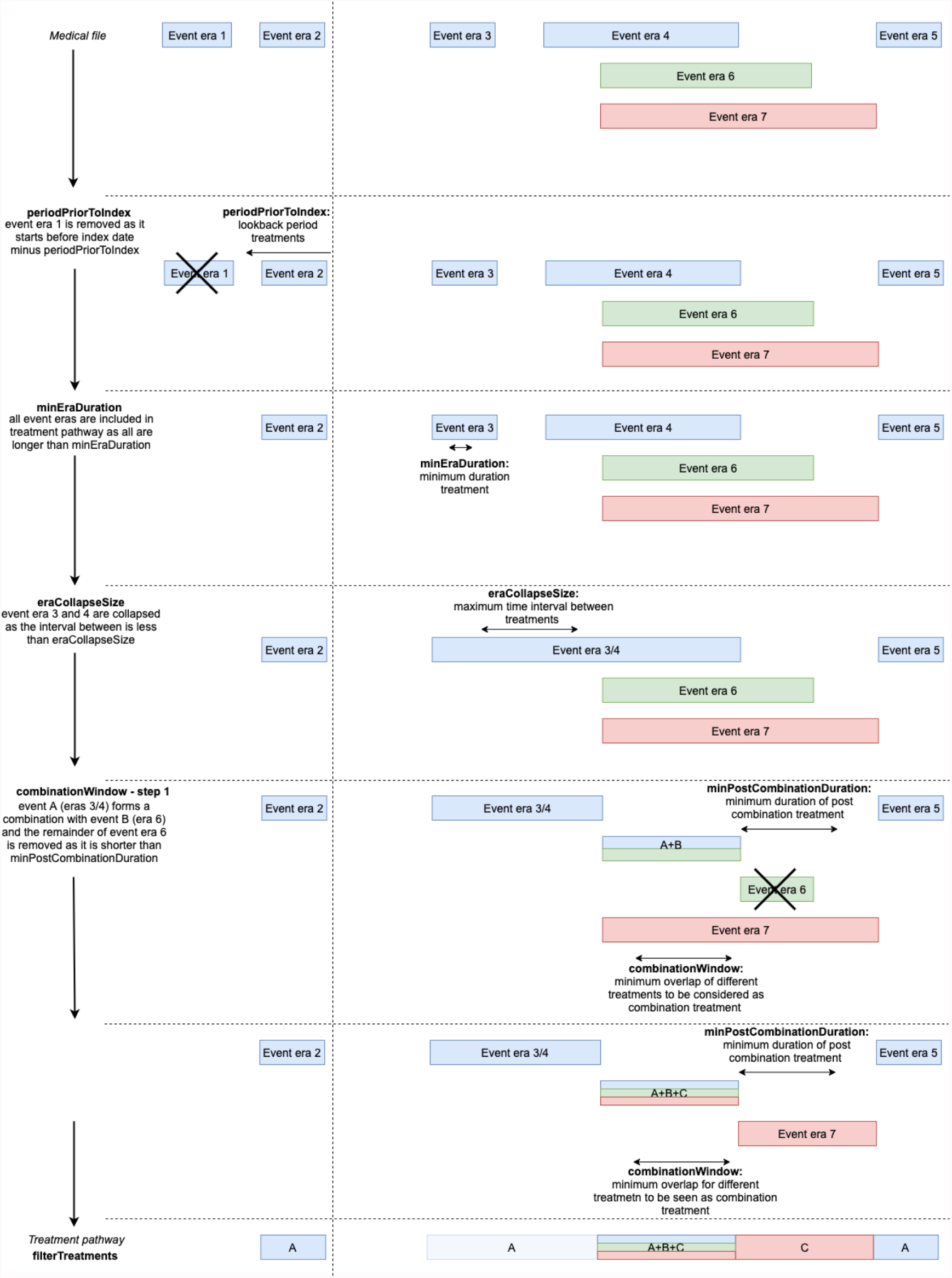

### Appendix B: Pathway settings

**Table 1:** List of pathway settings with description and expected input.

| General settings |  |  |
| --- | --- | --- |
| studyName | Unique name identifying the set of study parameters below | Name |
| targetCohortId | Select one study population | Cohort ID |
| eventCohortIds | Select all treatments of interest | List of cohort IDs |
| Individual pathway settings |  |  |
| periodPriorToIndex | The period (number of days) prior to the index date of the target cohort from which treatments should be included | Number of days |
| minEraDuration | Minimum time an event era should last to be included in the analysis | Number of days |
| eraCollapseSize | Maximum gap within two eras of the same event cohort which would still allow the eras to be collapsed into one era | Number of days |
| combinationWindow | Time that two event eras need to overlap to be considered a combination treatment | Number of days |
| minPostCombinationDuration | Minimum time that an event era before or after a generated combination treatment should last to be included in the pathway as a separate treatment | Number of days |
| filterTreatments | Select which treatments should be included in pathway first time occurrences of treatments ('First'), remove sequential repeated treatments ('Changes'), all treatments ('All') | First / Changes / All |
| maxPathLength | Maximum number of treatments included in pathway | Number of steps (max. 5) |

| Aggregate pathway settings |  |  |
| --- | --- | --- |
| minCellCount | Minimum number of persons with a specific treatment pathway for the pathway to be included in analysis | Number of subject |
| minCellMethod | Select to completely remove / sequentially adjust (by removing last step as often as necessary) treatment pathways below minCellCount | Remove / Adjust |
| groupCombinations | Select to group all non-fixed combinations in one category 'other' in the sunburst plot | TRUE / # below which to group |
| addNoPaths | Select to include untreated persons without treatment pathway in the sunburst plot | TRUE / FALSE |

### Appendix C: Example study type II diabetes mellitus, hypertension, and depression

#### Target cohorts

##### i) T2DM12mo

Initial Event Cohort:

People with continuous observation of 365 days before and 1,095 days after event may enter the cohort when observing any of the following:

- Condition occurrence of Diabetes* for the first time in the person’s history.

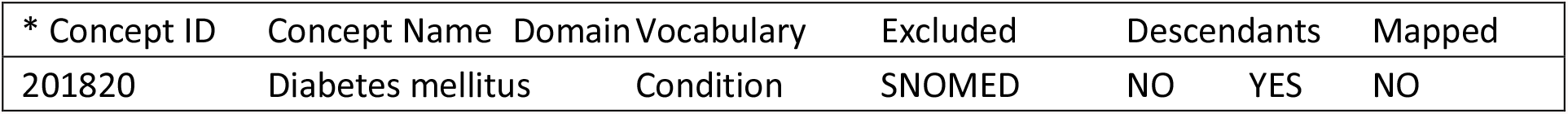

Inclusion Criteria:

- Age >= 18 years.
- No condition occurrences of Diabetes type 1** in their history or during follow-up time.

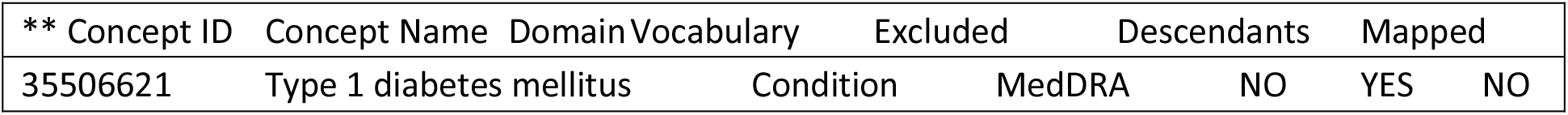

- Index date starting after December 31, 2009.

Cohort Exit: the person exits the cohort at the end of continuous observation.

##### ii) HTN12mo

Initial Event Cohort:

People with continuous observation of 365 days before and 1,095 days after event may enter the cohort when observing any of the following:

- Condition occurrence of Hypertension* for the first time in the person’s history.

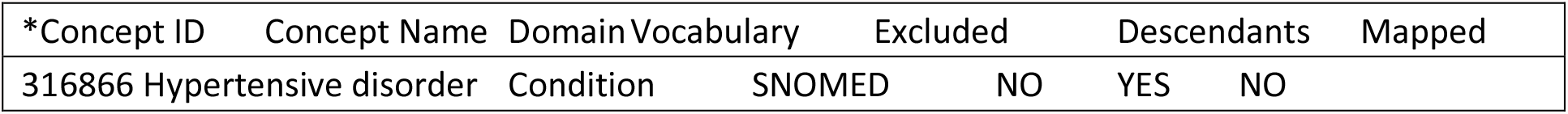

Inclusion Criteria:

- Age >= 18 years.
- Index date starting after December 31, 2009.

Cohort Exit: the person exits the cohort at the end of continuous observation.

##### iii) Depression12mo

Initial Event Cohort:

People with continuous observation of 365 days before and 1,095 days after event may enter the cohort when observing any of the following:

- Condition occurrence of Depression* for the first time in the person’s history.

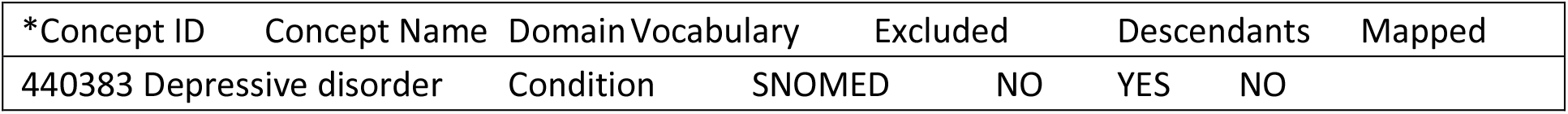

Inclusion Criteria:

- Age >= 18 years.
- No condition occurrences of Schizofrenia/Bipolar** in their history or during follow-up time.

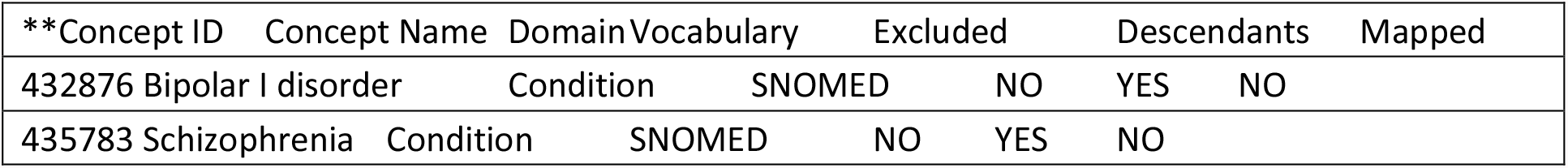

- Index date starting after December 31, 2009.

Cohort Exit: the person exits the cohort at the end of continuous observation.

#### Event cohorts

Initial Event Cohort:

People with continuous observation of 0 days before and 0 days after event may enter the cohort when observing any of the following:

- Drug exposure of treatment of interest*.

Cohort Exit: If the index event is found within an era, the cohort end date will use the era’s end date. Otherwise, it will use the observation period end date that contains the index event.

* Treatments of interest are defined by the following concept sets, details of Concept IDs can be found on https://athena.ohdsi.org/:

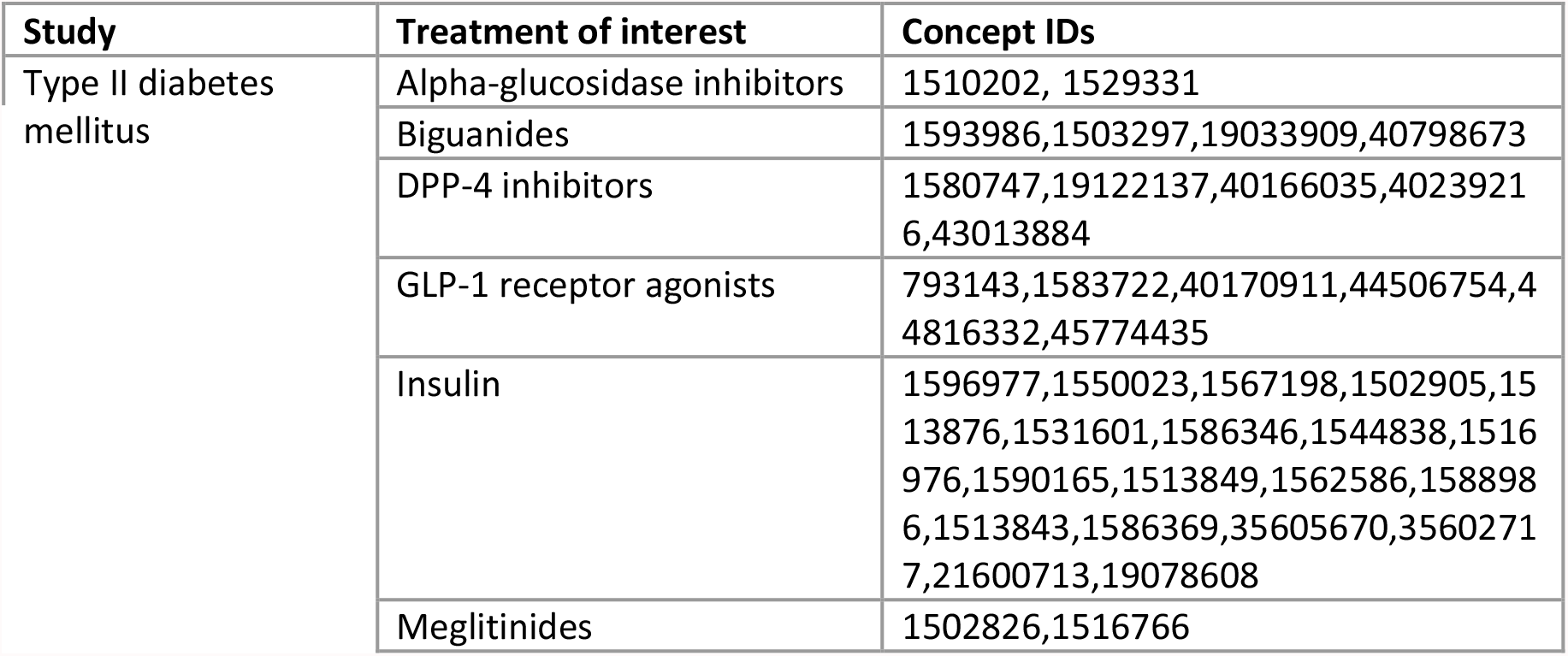
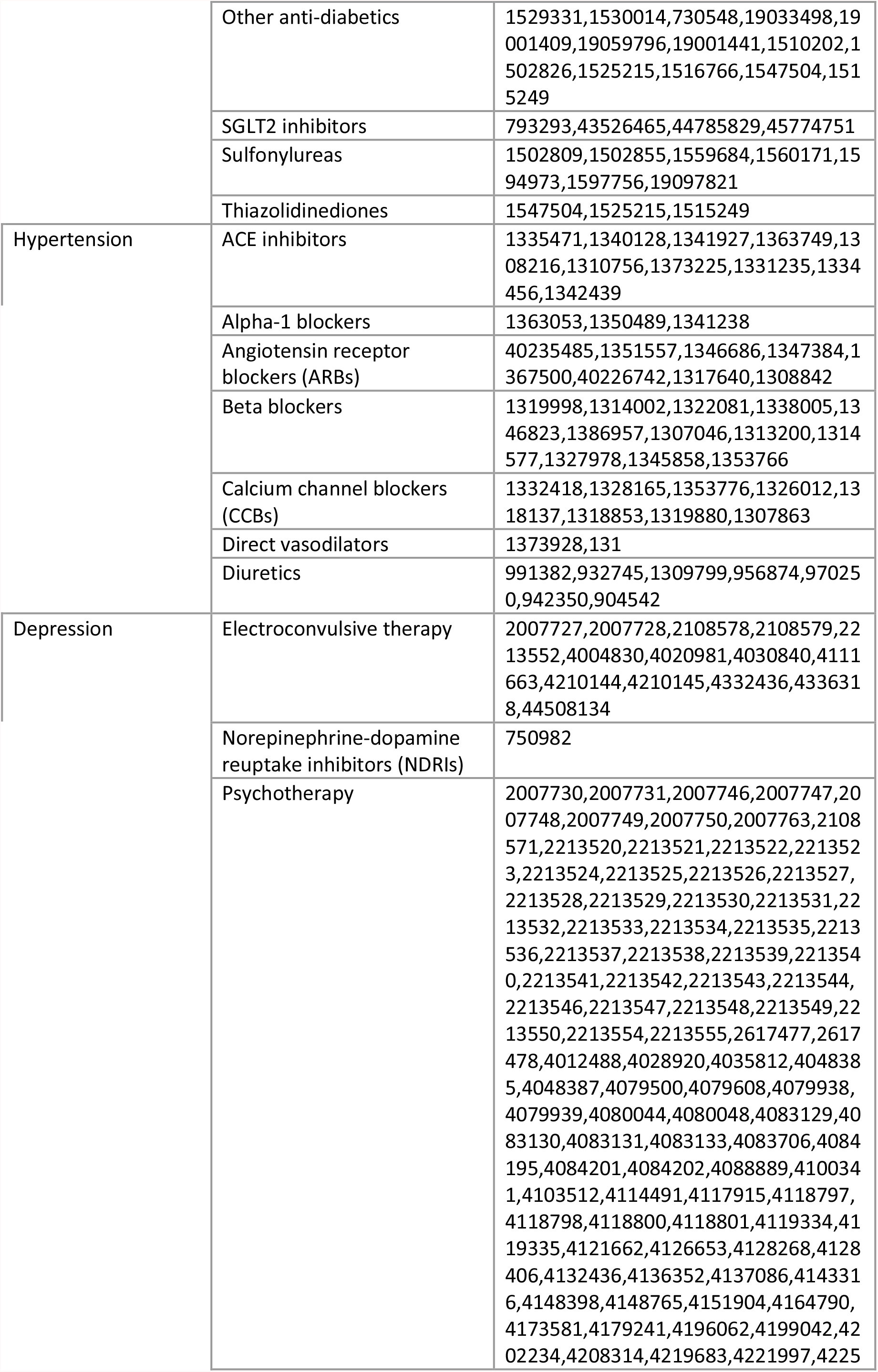
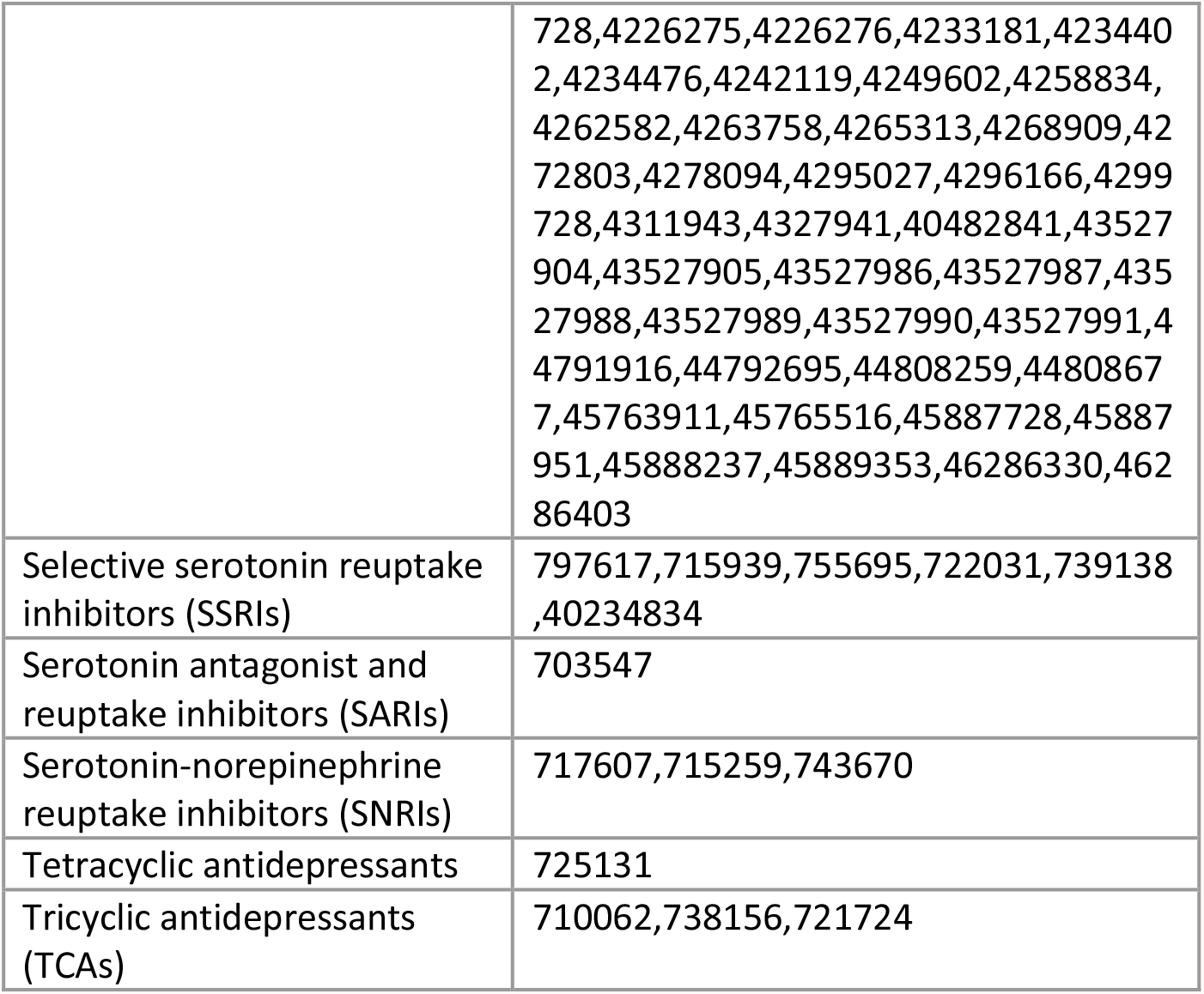

#### Baseline characterization

Baseline characterization:
| Covariate Name | Covariate ID (= Concept ID + time window 209) |
| --- | --- |
| Age | 1002 |
| Charlson comorbidity index score | 1901 |
| Male | 8507001 |
| Kidney disease | 198124209 |
| Diabetes mellitus | 201820209 |
| Heart failure | 316139209 |
| Hypertensive disorder | 316866209 |
| Cerebrovascular disease | 381591209 |
| Obesity | 433736209 |
| Depressive disorder | 440383209 |
| Ischemic heart disease | 4185932209 |

## Appendix D: Code to run example treatment patterns study

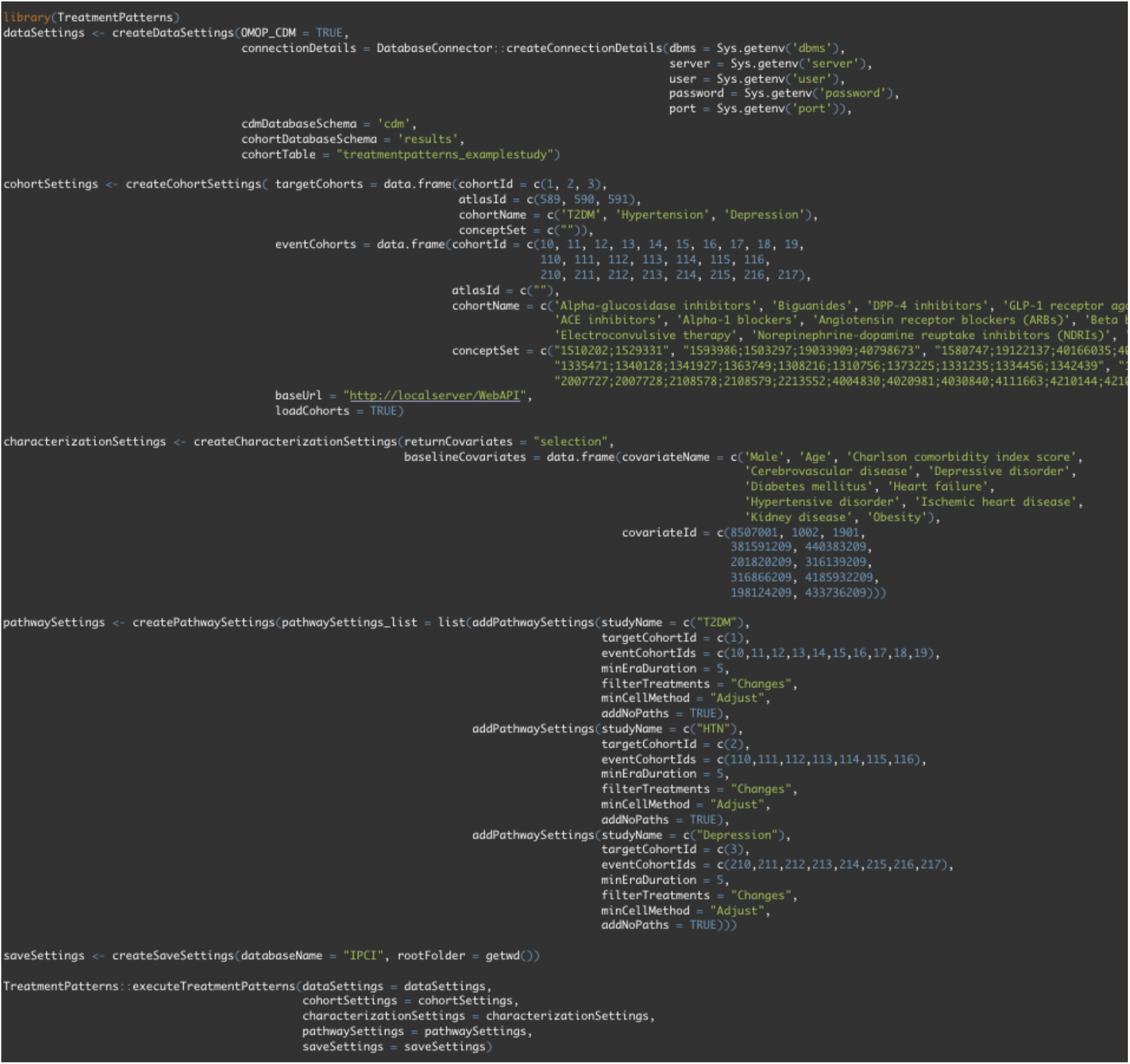

## Appendix E: Results example treatment patterns study

**Figure 1:**
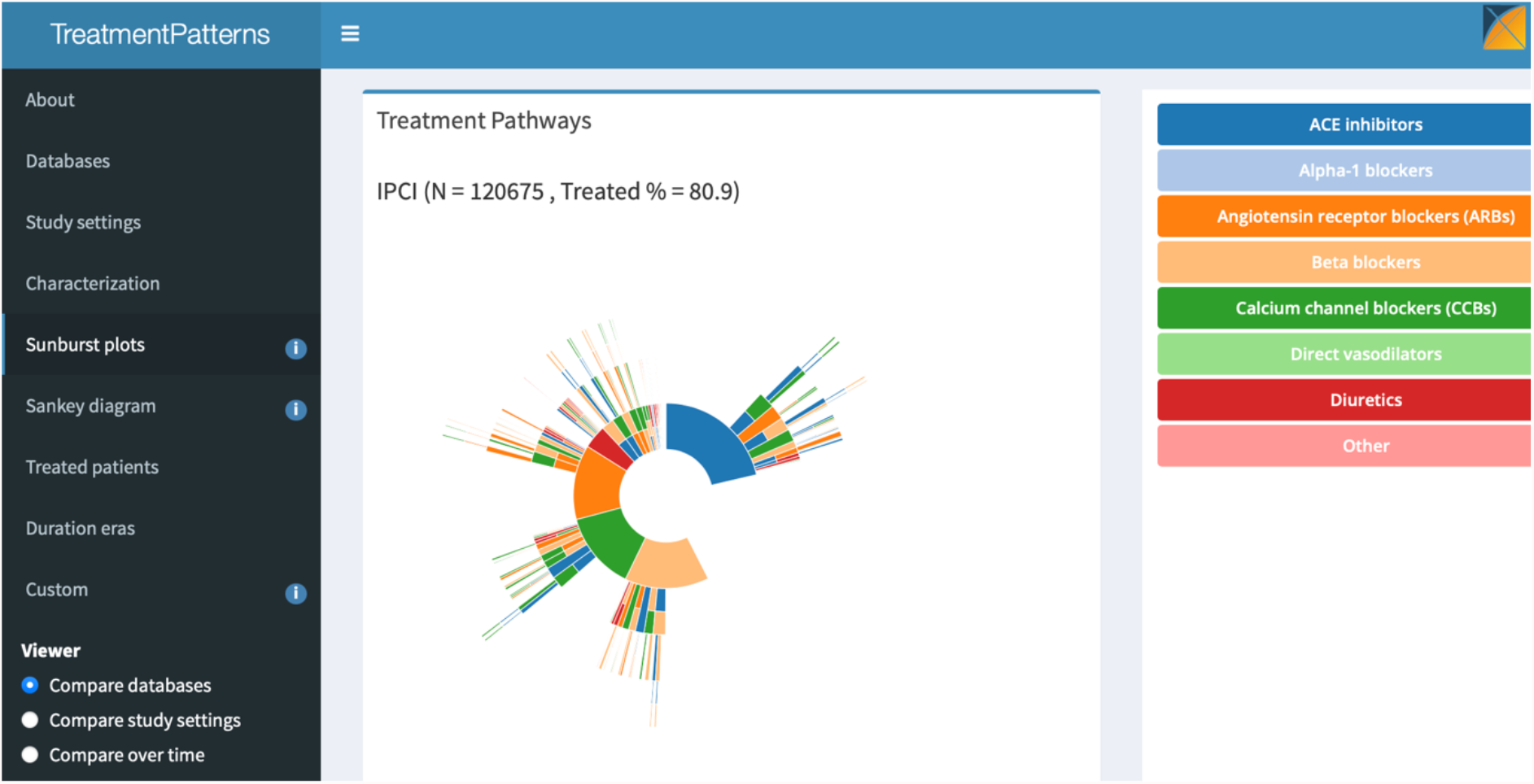
Sunburst plot visualizing the treatment pathways of patients with hypertension in IPCI.

**Figure 2:**
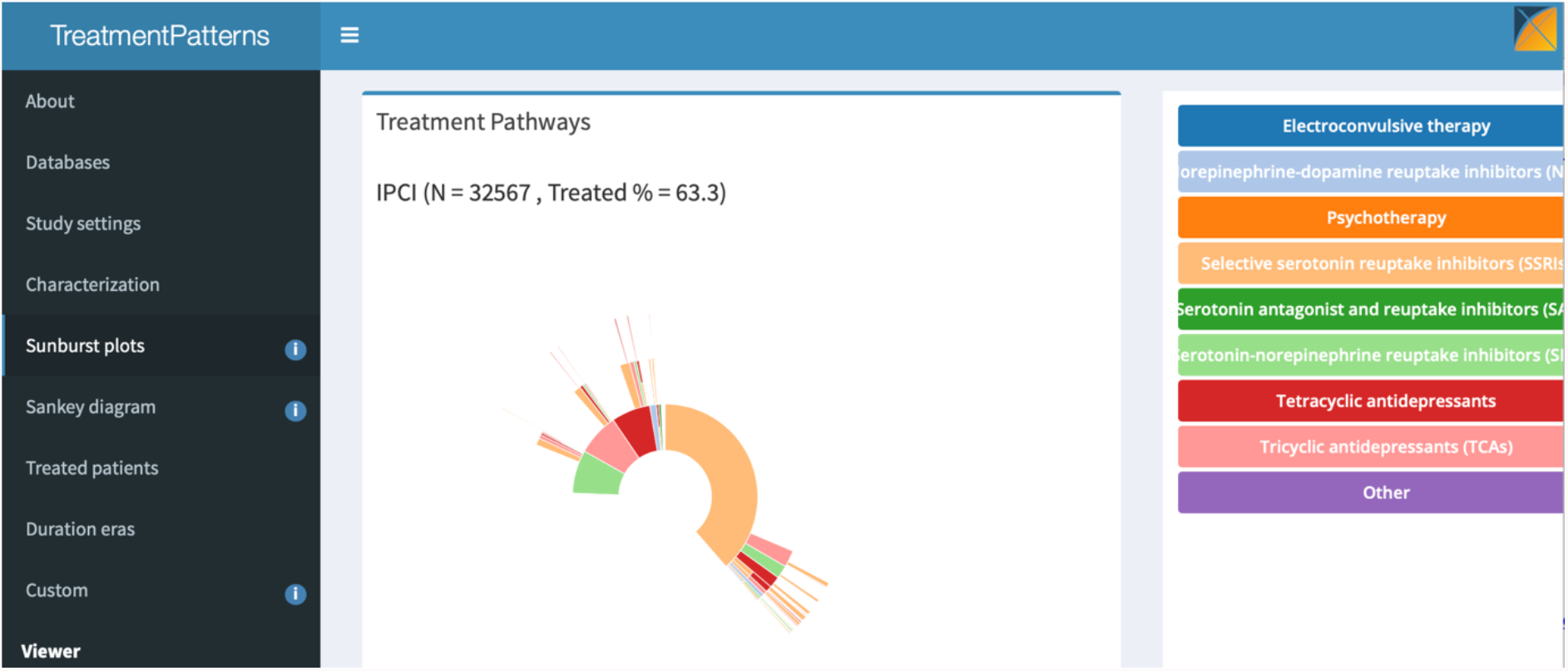
Sunburst plot visualizing the treatment pathways of patients with depression in IPCI.

**Figure 3:**
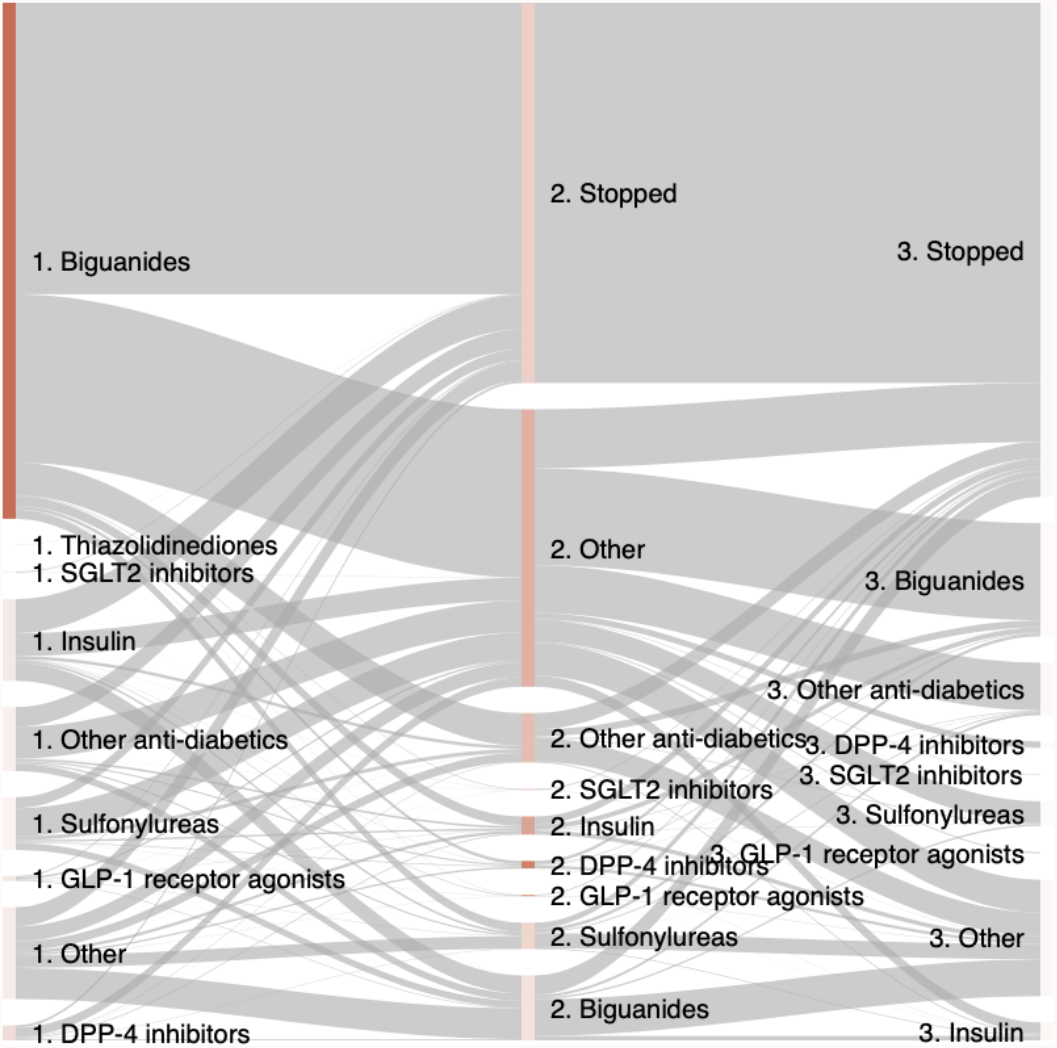
Sankey diagram visualizing the treatment pathways of patients with type II diabetes mellitus in IPCI.

**Figure 4:**
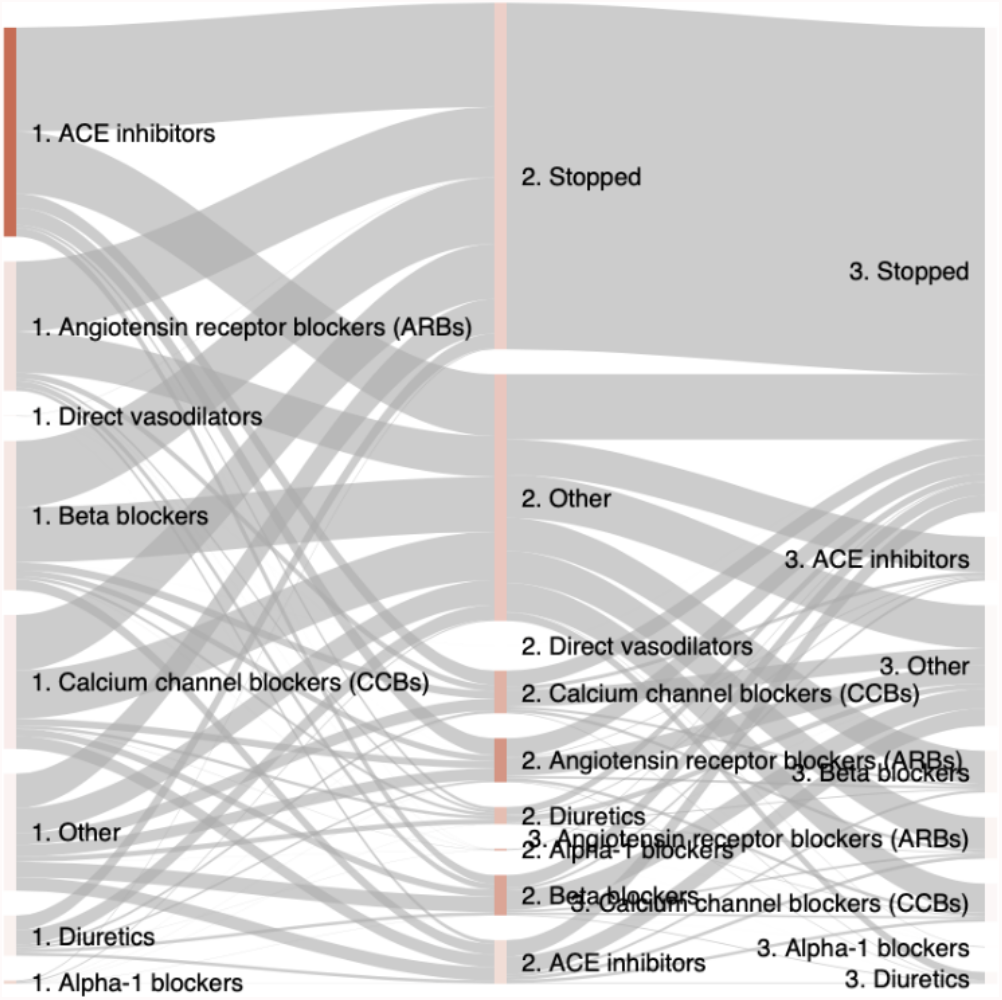
Sankey diagram visualizing the treatment pathways of patients with hypertension in IPCI.

**Figure 5:**
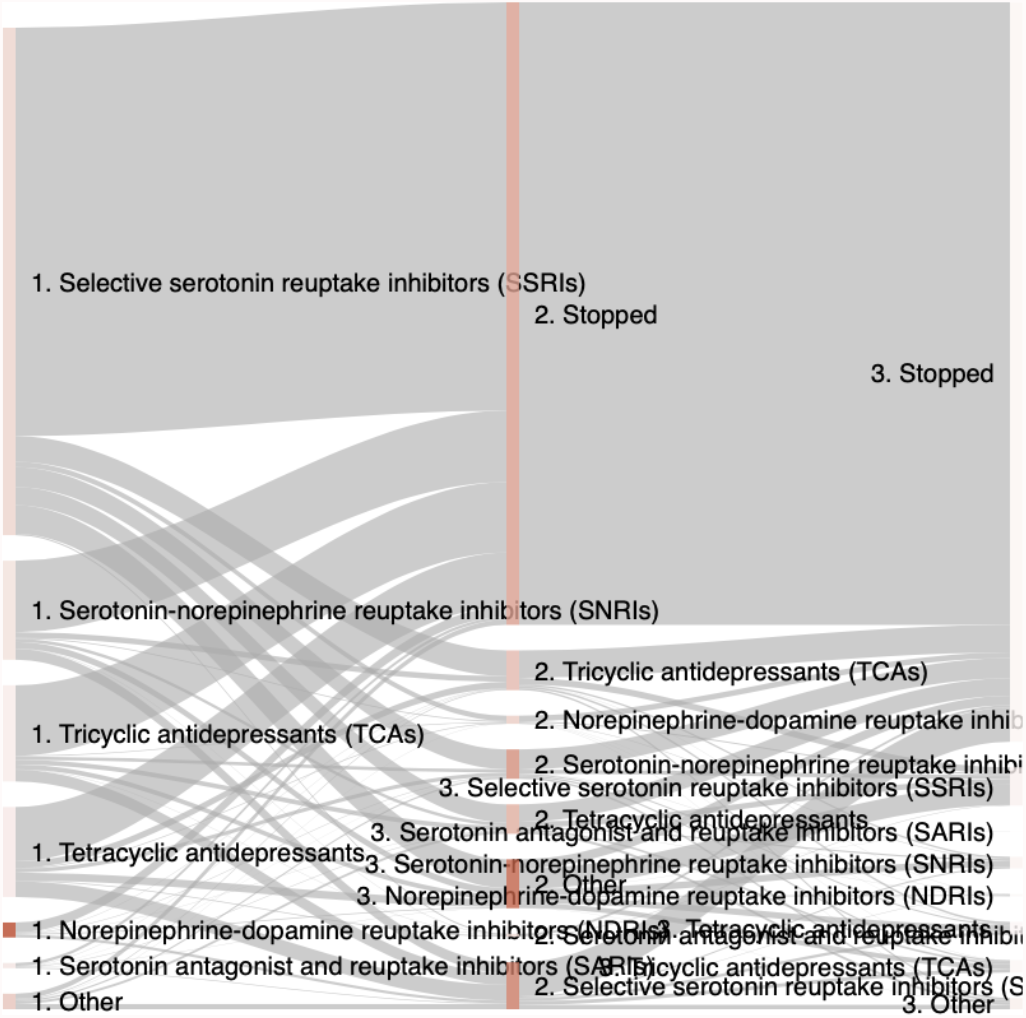
Sankey diagram visualizing the treatment pathways of patients with depression in IPCI.

**Figure 6:**
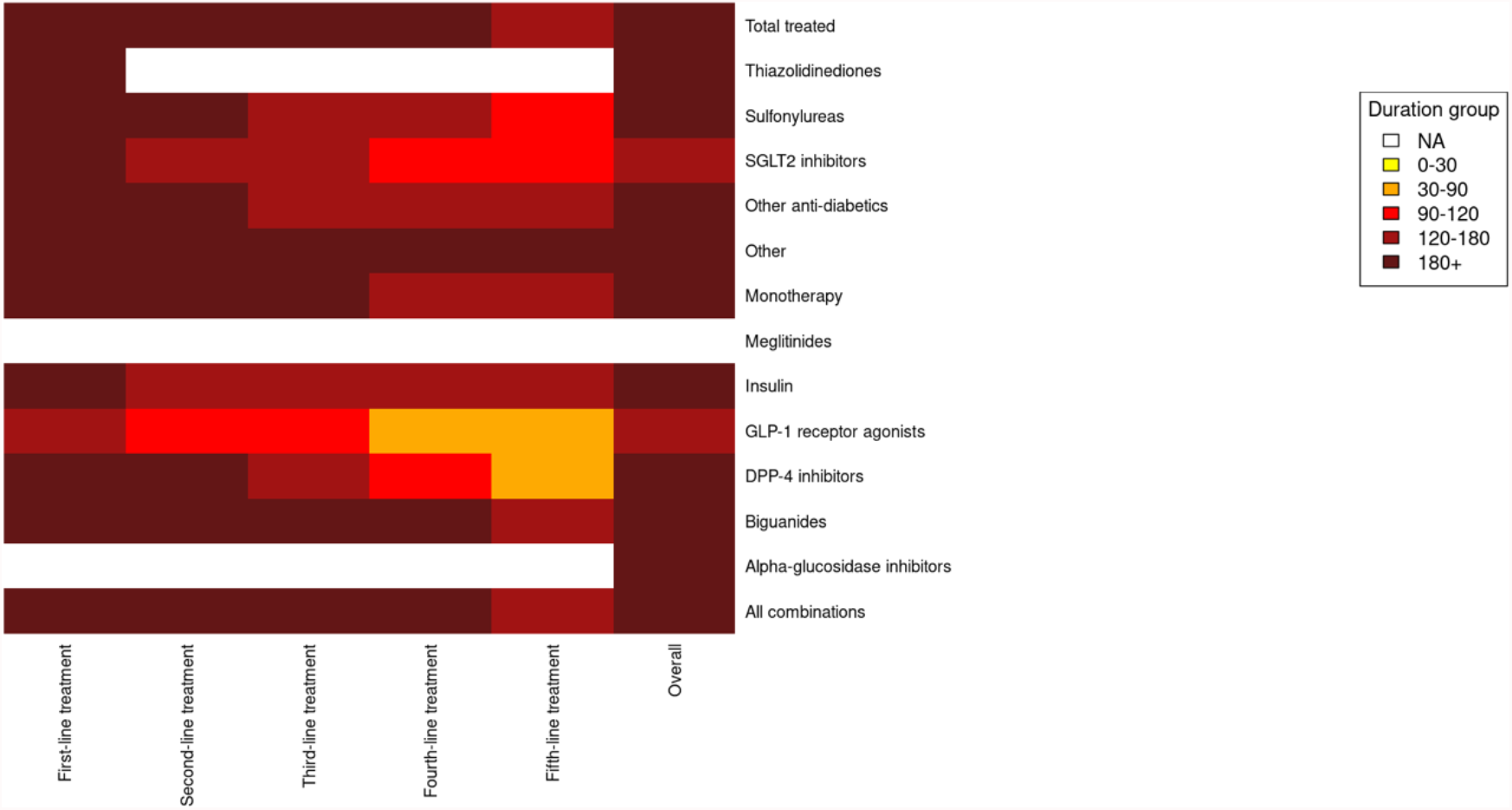
Heatmap visualizing duration of treatments of patients with type II diabetes mellitus in IPCI.

**Figure 7:**
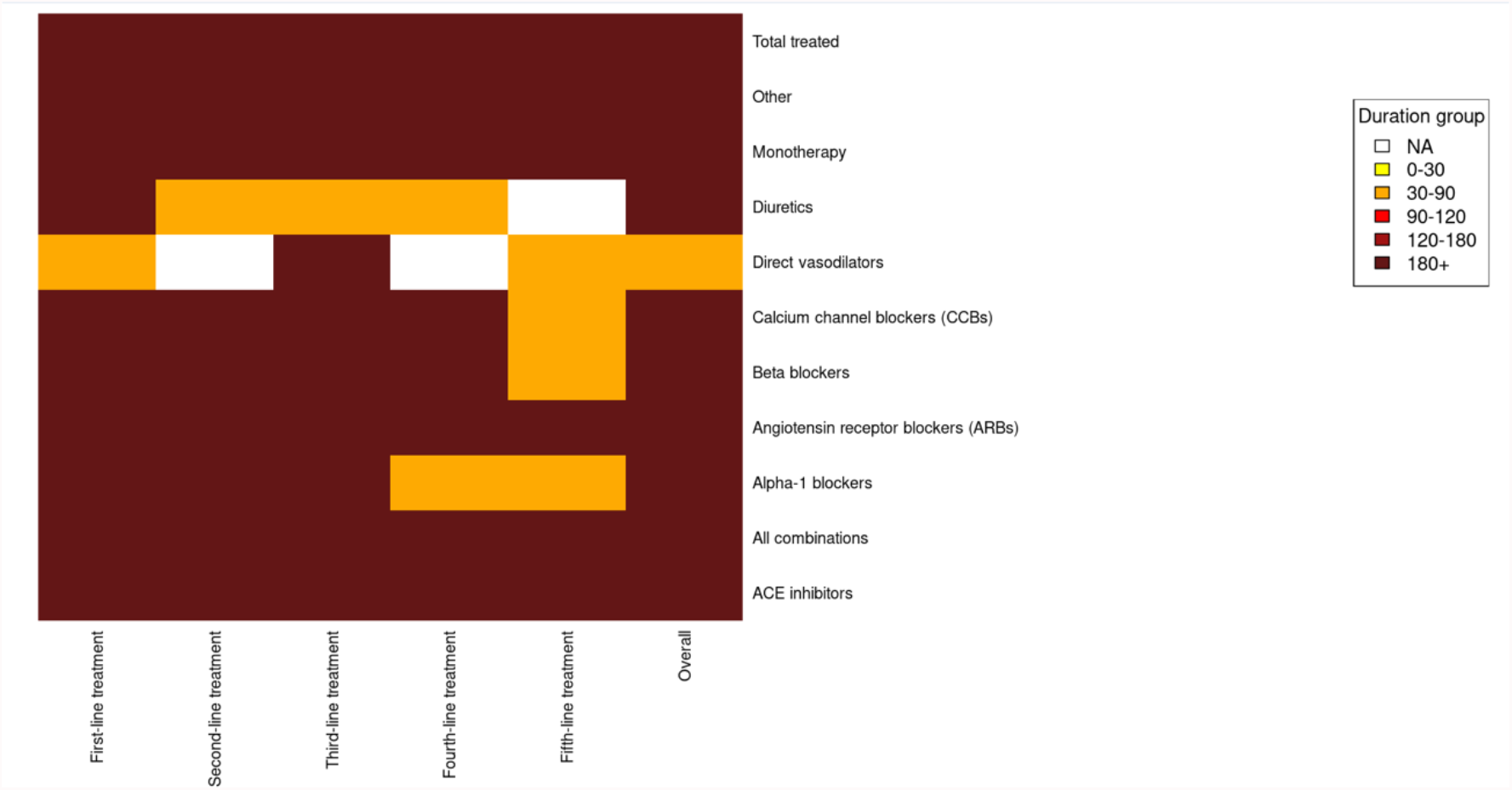
Heatmap visualizing duration of treatments of patients with hypertension in IPCI.

**Figure 8:**
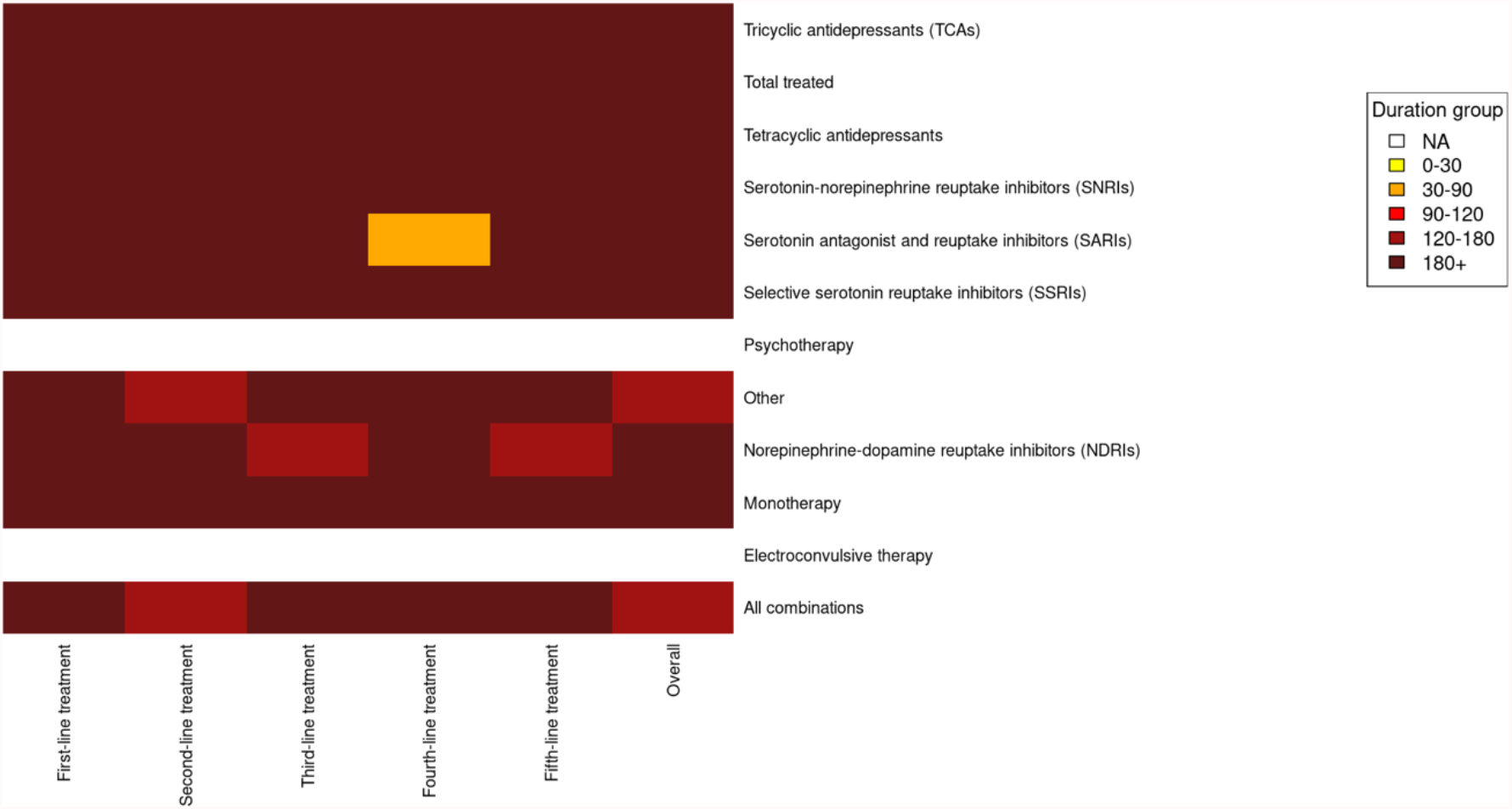
Heatmap visualizing duration of treatments of patients with depression in IPCI.

## Footnotes

1 https://github.com/OHDSI/Hades

2 https://www.ipci.nl/

3 Available in ATLAS: https://atlas-demo.ohdsi.org.

